# High SARS-CoV-2 viral load is associated with a worse clinical outcome of COVID-19 disease

**DOI:** 10.1101/2020.11.13.20229666

**Authors:** María Eugenia Soria, Marta Cortón, Brenda Martínez-González, Rebeca Lobo-Vega, Lucía Vázquez-Sirvent, Rosario López-Rodríguez, Berta Almoguera, Ignacio Mahillo, Pablo Mínguez, Antonio Herrero, Juan Carlos Taracido, Alicia Macías-Valcayo, Jaime Esteban, Ricardo Fernandez-Roblas, Ignacio Gadea, Javier Ruíz-Hornillos, Carmen Ayuso, Celia Perales

**Author notes:** These authors have contributed equally to this work (María Eugenia Soria, Marta Cortón). <u>Associated clinical group of University Hospital Fundación Jiménez Díaz:</u>. Miguel Górgolas, Alfonso Cabello, Germán Peces Barba, Sara Heili, César Calvo, M^a^ Dolores Martín Ríos, Arnoldo Santos, Olga Sánchez-Pernaute, Lucía Llanos, Sandra Zazo, Federico Rojo, Felipe Villar, Raimundo de Andrés, Ignacio Jiménez Alfaro.

## Abstract

COVID-19 severity and progression are determined by several host and virological factors that may influence the final outcome of SARS-CoV-2-infected patients. The objective of this work is to determine a possible association between the viral load, obtained from nasopharyngeal swabs, and the severity of the infection in a cohort of 448 SARS-CoV-2-infected patients from a hospital in Madrid during the first outbreak of the pandemic in Spain. To perform this, we have clinically classified patients as mild, moderate and severe COVID-19 according to a number of clinical parameters such as hospitalization requirement, need of oxygen therapy, admission to intensive care units and/or exitus. Here we report a statistically significant correlation between viral load and disease severity, being high viral load associated with worse clinical prognosis, independently of several previously identified risk factors such as age, sex, hypertension, cardiovascular disease, diabetes, obesity, and lung disease (asthma and chronic obstructive pulmonary disease). The data presented here reinforce the viral load as a potential biomarker for predicting disease severity in SARS-CoV-2-infected patients. It is also an important parameter in viral evolution since it relates to the numbers and types of variant genomes present in a viral population, a potential determinant of disease progression.

## Introduction

Coronavirus SARS-CoV-2 emerged in the human population in 2019 is the causal agent of the new pandemic disease COVID-19 (1). The virus has spread rapidly worldwide, and at the time of this writing there are 46,403,652 confirmed cases, and 1,198,569 deaths in 219 countries worldwide, according to the WHO (https://covid19.who.int/); these numbers are increasing daily. Evolution of a virus in a specific host is defined by a number of closely related parameters such as viral load, replication rate, genetic heterogeneity and viral fitness that may influence virus adaptability, viral pathogenesis and disease progression (2, 3). The replicative capacity of a virus is clinically relevant because it largely determines the viral load in infected individuals, and viral load influences disease manifestations (4). In the case of SARS-CoV-2, some studies have correlated viral load with disease severity whereas in others this correlation was not clear. A positive correlation was reported in a cohort of SARS-CoV-2-infected patients from China, showing that the viral load detected in the respiratory tract was positively linked to lung disease severity (5). In a related study, the analysis of the viral RNA level in upper respiratory tract samples from 76 patients with COVID-19 revealed significantly lower Ct values (cycle threshold which is inversely correlated with viral RNA level), and longer virus-shedding periods in those patients classified as severe, as compared with those that exhibited mild disease (6). Additionally, a prospective study in a large hospitalized cohort of 1,145 infected patients documented a significant lower probability of survival in patients with high viral load than in those with low viral load (7). In contrast, in other studies, it was observed that the diagnostic viral load level was lower in hospitalized than in non-hospitalized patients, resulting in a lack of correlation of viral load with admission to intensive care unit (ICU), length of oxygen support, and overall patient survival (8). Thus, the dynamics of viral load and its connection with different clinical parameters remain poorly characterized, and large studies with additional cohorts worldwide are needed to define the possible association and the predictive value of the viral load regarding disease progression and mortality. Viral RNA load calculated from nasopharyngeal swabs might be added to other predictive parameters to complete an early risk stratification of COVID-19 patients (9).

## Results and Discussion

Given the disparate results trying to correlate viral load with COVID-19 disease progression, we have addressed this question with a large cohort of 448 patients admitted to the Fundación Jiménez Díaz Hospital (FJD, Madrid, Spain) from April 3 to 29, 2020 coinciding with the first COVID-19 outbreak in Spain. At the time of admission, all patients had clinical symptoms, and were confirmed to be positive for SARS-CoV-2 by a specific real-time RT-PCR (VIASURE Real Time PCR). The Ct values analyzed correspond to the first available COVID-19 PCR-positive sample for each patient. Details regarding the clinical classification are described in Materials and Methods. Mean Ct values for mild, moderate and severe COVID-19 patients were 35.75 ± 0.45, 32.69 ± 0.37, and 29.58 ± 0.70, respectively. A univariate analysis showed statistically significant differences among viral load values according to the infection severity (p<0.0001; ANOVA test) (Figure 1). Specifically, average Ct values were significantly lower in the severe group as compared with the moderate and the mild disease groups, and also for the comparison between moderate and mild clinical categories (p<0.001 in all cases) (t-test with Bonferroni correction). Thus, Ct values (as a measurement of viral load) in SARS-CoV-2-infected patients correlates positively with the disease progression and poor prognosis.

**Figure 1.**
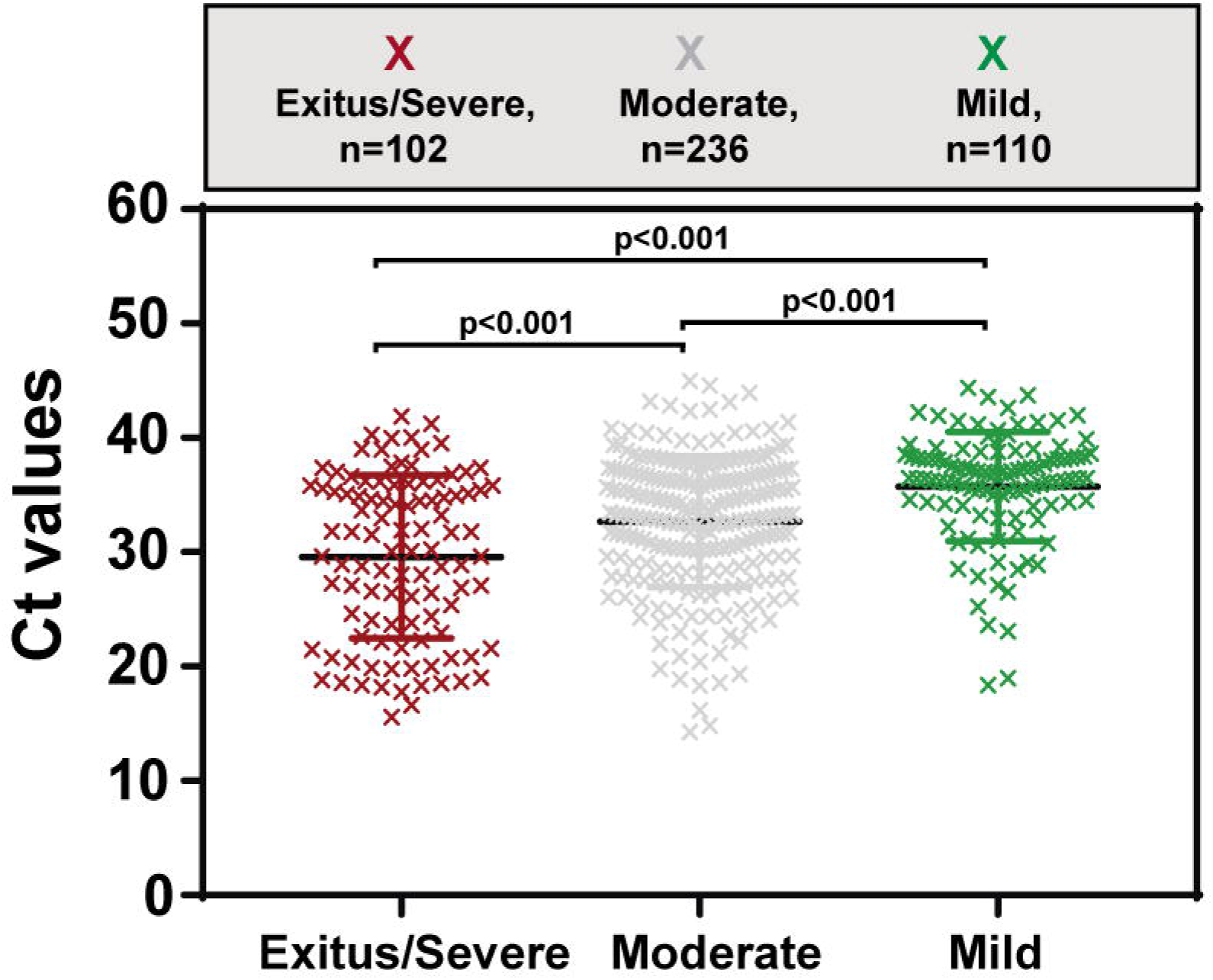
Correlation between viral load [measured by the Ct (number of PCR amplification cycles needed to cross the threshold detection level) value] and the severity of COVID-19 disease.

Regarding host factors, it has been established that age greater than 65 is a risk factor to develop acute respiratory distress syndrome (ARDS), a major complication of COVID-19 pneumonia, and that the risk of death increases with advanced age (10). As a second factor, disease severity and mortality for males is significantly higher than for females, being older men the population most at risk (11, 12). Additionally, several co-morbidities have been potentially associated with poor outcome including hypertension (high blood pressure), cardiovascular disease, diabetes, obesity, and lung disease (asthma, EPOC) (13). We have included these risk factor data in our patient cohort to assess their alignment with our disease severity-viral RNA load correlation. Interestingly, a significant association between viral load and infection severity was still observed after adjusting for age, sex, hypertension, cardiovascular disease, diabetes, obesity, asthma and chronic obstructive pulmonary disease (COPD) (see ANCOVA tests in Table 1). The significant difference in viral load between the three groups was not attributable to the percentage of hospitalization, the percentage of ICU admission or the percentage of the different types of oxigenotherapy (see ANCOVA tests in Table 1). Thus, our results with a SARS-CoV-2 population from a cohort which is different from those in previous studies on viral load influence on disease, suggest a positive correlation between viral load and COVID-19 disease severity. This conclusion is also in agreement with two studies showing a longer persistence of high viral load viruses in respiratory samples of patients with severe disease than those with mild disease, suggesting that the viral load may be a prognostic parameter (14, 15).

**Table 1.** Demographic data and preexisting comorbidities in 448 SARS-CoV-2-infected patients classified by disease severity of COVID-19.

| Characteristics | Total<br>(n=448) | Disease Severity |  |  | p-value<sup>a</sup> | Significance<sup>a</sup> |
| --- | --- | --- | --- | --- | --- | --- |
|  |  | Exitus/Severe<br>(n=102) | Moderate<br>(n=236) | Mild<br>(n=110) |  |  |
| Age mean ± standard deviation, years | 71.04 ± 18.29 | 79.92 ± 15.33 | 73.25 ± 16.59 | 58.05 ± 17.50 | 1.49 x 10<sup>-12</sup> | *** |
| Age > 60 years (%) | 321 (71.6%) | 91 (89.2%) | 185 (78.4%) | 45 (41.0%) | 1.11 x 10<sup>-8</sup> | *** |
| Male (%) | 205 (45.7%) | 49 (48.0%) | 122 (51.7%) | 34 (31.0%) | 1.40 x 10<sup>-12</sup> | *** |
| Female (%) | 243 (54.2%) | 53 (52.0%) | 114 (48.3%) | 76 (69.1%) |  |  |
| Hypertension (%) | 236 (52.7%) | 69 (67.6%) | 134 (56.8%) | 33 (30.0%) | 1.14 x 10<sup>-12</sup> | *** |
| Cardiac disease (%) | 142 (31.7%) | 42 (41.2%) | 73 (30.9%) | 27 (24.5%) | 1.35 x 10<sup>-12</sup> | *** |
| Diabetes (%) | 8 (1.8%) | 2 (2.0%) | 5 (2.1%) | 1 (0.9%) | 1.08 x 10<sup>-12</sup> | *** |
| Obesity (%) | 17 (3.8%) | 5 (4.9%) | 8 (3.4%) | 4 (3.6%) | 1.37 x 10<sup>-12</sup> | *** |
| Asthma (%) | 29 (6.5%) | 8 (7.8%) | 16 (6.8%) | 5 (4.5%) | 1.42 x 10<sup>-12</sup> | *** |
| COPD (%) | 15 (3.3%) | 4 (3.9%) | 8 (3.4%) | 3 (2.7%) | 1.34 x 10<sup>-12</sup> | *** |
| Hospitalization (%)<sup>b</sup> | 365 (81.5%) | 102 (100%) | 236 (100%) | 27 (24.5%) | 1.21 x 10<sup>-12</sup> | *** |
| ICU admission (%)<sup>c</sup> | 36 (8.0%) | 33 (32.3%) | 2 (0.8%) | 1 (0.9%) | 1.16 x 10<sup>-12</sup> | *** |
| Conventional oxygen therapy (%) | 288 (64.3%) | 102 (100%) | 183 (77.5%) | 3 (2.7%) | 1.10 x 10<sup>-12</sup> | *** |
| Invasive mechanical ventilation (%) | 19 (4.2%) | 19 (18.6%) | 0 (0%) | 0 (0%) | 1.25 x 10<sup>-12</sup> | *** |
| Noninvasive mechanical ventilation (%)<sup>d</sup> | 21 (4.7%) | 20 (19.6%) | 1 (0.4%) | 0 (0%) | 1.27 x 10<sup>-12</sup> | *** |
| High Flow Nasal Cannulas (%) | 19 (4.2%) | 19 (18.6%) | 0 (0%) | 0 (0%) | 1.43 x 10<sup>-12</sup> | *** |
| Survival 90 days after diagnosis (%)<sup>e</sup> | 366 (81.7%) | 27 (26.5%) | 233 (98.7%) | 106 (96.4%) | 8.44 x 10<sup>-13</sup> | *** |
| Days since symptoms onset<sup>f</sup> | 7.84 ± 6.40 | 7.57 ± 6.62 | 8.26 ± 6.53 | 3.25 ± 0.50 | 2.81 x 10<sup>-12</sup> | *** |

<sup>a</sup>p-values indicate correlation between viral load and disease severity after adjusting for the baseline characteristics, pre-existing comorbidities, hospitalization, oxygen therapy and mortality. Statistical significance: <sup>b</sup> $P < 0.001$ ; ANCOVA test.
<sup>b</sup>Exceptions are the following: the 27th patients classified as mild COVID-19 were hospitalized for causes other than SARS-CoV-2 infection. In the group moderate COVID-19, one patient was hospitalized for causes other than SARS-CoV-2 infection, but was assigned to this group due COVID-19 pneumonia according to the clinical history.
<sup>c</sup>Exceptions are the following: two patients in the group moderate COVID-19 and one patient in the group mild COVID-19 were admitted to ICU for causes other than SARS-CoV-2 infection.
<sup>d</sup>Exceptions are the following: in the group moderate COVID-19, one patient required noninvasive mechanical ventilation during hospitalization due to a previous pathology.
<sup>e</sup>Exceptions are the following: three patients in the group moderate COVID-19 and four patients in the group mild COVID-19 died during the 90 days after diagnosis due to pathologies prior to SARS-CoV-2 infection.
<sup>f</sup>Data available for 102 patients.

To monitor the average number of days between the symptom onset and the sample extraction in our cohort, we collected these data for 102 patients. On average NP swabs were obtained at 7.84 ± 6.40 days after symptoms onset, which is within the time interval of active infection. It has been described that the highest SARS-CoV-2 viral load in throat swabs -and consequently the highest transmissibility peaks- is around 5-6 days after the symptom onset (16), but this is an average that depends on each patient; the transmissibility window extends from a few days before symptom onset up to 30 days in patients with severe disease (14, 17).

A correlation between high viral load and disease severity has been also found in other viral infections. Among children naturally infected with respiratory syncytial virus (RSV), increased viral load was associated with clinical severity of disease defined as an increased risk for intensive care, prolonged hospitalization, or the development of respiratory failure (18). Examination of hepatitis A virus (HAV) RNA from sera by real-time PCR resulted in higher initial viral load in patients with severe outcomes such as fulminant hepatitis and severe acute hepatitis than in patients with less severe infection (19). Viral load in patients infected with the pandemic type A influenza virus H1N1 (2009) who suffered pneumonia was higher than in patients with milder disease (those with bronchitis or upper respiratory tract infection), suggesting that the viral load is also an important predictive value in influenza infection (20). Other instances of a connection of viral load with disease progression have been reviewed (2).

In terms of viral quasispecies dynamics, a key question regarding clinical implications is that a population with a large viral load will contain a broader mutant repertoire than a population with low viral load. Even if each individual mutant might be present at the same frequency in both populations (as it corresponds to mutant frequency being an “intrinsic” property), the mutant repertoire is an “extrinsic” property of the population, a key distinction that has been numerically studied (21). Thus, viral load is a potential source of variants that enhance the probability of infection to alternative cell types in the course of infection which is increasingly viewed as a step-wise adaptation process (2-4, 21).

## Methods

### Patient cohort and stratification

Data collected included patient demographics, risk factors for SARS-CoV-2 disease and clinical information related to the time of SARS-CoV-2 diagnosis (Table 1). Patients were classified according to the following COVID-19-associated parameters: (1) need of hospital admission, (2) need for mechanical ventilation, (3) admission to the ICU, and (4) exitus attributed to COVID-19. Patients were classified as mild, moderate and severe cases according to the requirement and the type of hospitalization: (1) mild symptoms (neither hospital admission nor ICU) (n=110), (2) moderate symptoms (hospitalization without ICU) (n=236), and (3) severe symptoms (hospitalization with admission to the ICU, and/or exitus) (n=102). Exceptions to these criteria are detailed in Table 1. The clinical relevance was defined before the data analysis was performed.

### Molecular testing for SARS-CoV-2 by Ct value measurements

Nasopharyngeal swabs were collected at FJD hospital by trained medical personnel from all patients included in the study due to suspected COVID-19 infection. After collection, the NP samples were transferred to viral transport media and transported to the Microbiology Department for molecular testing. RT-PCR to obtain diagnostic SARS-CoV-2 viral load was performed using the kit VIASURE Real Time PCR Detection Kits by CerTest BIOTEC following the manufacturer instructions. The Ct values were calculated using SARS-CoV-2-specific oligonucleotides directed to the ORF1ab.

### Statistics

The statistically significance of differences among viral load values according to the infection severity was calculated with the ANOVA test and t-test with Bonferroni correction using GraphPad Prism 7.00. The association between viral load and disease severity adjusted by risk factors was calculated with ANCOVA test using software R version 4.0.2.

### Study approval

All samples were collected according to WHO guidelines. This study was approved by the Ethics Committee and the Institutional Review Board of the FJD hospital (no. PIC-087-20-FJD).

## Data Availability

All manuscript data will be available upon request.

## Authors contributions

CP, IG, RF-R and JE designed the study. AM-V, IG, RF-R and JE collected samples and validated RT-PCRs. MES, BM-G, RL-V and LV-S analyzed Ct and clinical data. MC, RL-R, BA, PM, AH, JCT, JR-H, CA and CP classified clinically SARS-CoV-2 patients. IM performed statistics. JMA, JR-H, CA and CP supervised analysis. CP, MES, BM-G, RL-V and LV-S wrote the manuscript. All authors reviewed the manuscript.

## Acknowledgements

We acknowledge all people in the Clinical Microbiology Department of the FJD for helping with the sample and data collection. We thank all health-care professionals who attended COVID-19 patients and collected the clinical samples that were included in this study in a difficult moment of the COVID-19 epidemic in Spain. We thank José María Aguado and Octavio Carretero for their support in the whole project. We are indebted to E. Domingo for encouragement and critical reading of the manuscript, and to Nuria Verdaguer and Enrique Marcos for valuable discussions about SARS-CoV-2 scientific findings. This work was supported by Instituto de Salud Carlos III, Spanish Ministry of Science and Innovation (COVID-19 Research Call COV20/00181) — co□financed by European Development Regional Fund “A way to achieve Europe”. The work was also supported by grants CSIC-COV19-014 from Consejo Superior de Investigaciones Científicas (CSIC), BFU2017-91384-EXP from Ministerio de Ciencia, Innovación y Universidades (MICIU), PI18/00210 from Instituto de Salud Carlos III. C.P., M.C. and P.M. are supported by the Miguel Servet program of the Instituto de Salud Carlos III (CPII19/00001, CPII17/00006 and CP16/00116, respectively) cofinanced by the European Regional Development Fund (ERDF). CIBERehd (Centro de Investigación en Red de Enfermedades Hepáticas y Digestivas) is funded by Instituto de Salud Carlos III. Institutional grants from the Fundación Ramón Areces and Banco Santander to the CBMSO are also acknowledged. The team at CBMSO belongs to the Global Virus Network (GVN). B. M.-G. is supported by predoctoral contract PFIS FI19/00119 from Instituto de Salud Carlos III (Ministerio de Sanidad y Consumo) cofinanced by Fondo Social Europeo (FSE). R. L.-V. is supported by predoctoral contract PEJD-2019-PRE/BMD-16414 from Comunidad de Madrid. R L-R is sponsored by the IIS-Fundación Jiménez Díaz-UAM Genomic Medicine Chair.

